# Development of an amplicon-based sequencing approach in response to the global emergence of human monkeypox virus

**DOI:** 10.1101/2022.10.14.22280783

**Authors:** Nicholas F.G. Chen, Chrispin Chaguza, Luc Gagne, Matthew Doucette, Sandra Smole, Erika Buzby, Joshua Hall, Stephanie Ash, Rachel Harrington, Seana Cofsky, Selina Clancy, Curtis J. Kapsak, Joel Sevinsky, Kevin Libuit, Daniel J. Park, Peera Hemarajata, Jacob M. Garrigues, Nicole M. Green, Sean Sierra-Patev, Kristin Carpenter-Azevedo, Richard C. Huard, Claire Pearson, Kutluhan Incekara, Christina Nishimura, Jian Ping Huang, Emily Gagnon, Ethan Reever, Jafar Razeq, Anthony Muyombwe, Vítor Borges, Rita Ferreira, Daniel Sobral, Silvia Duarte, Daniela Santos, Luís Vieira, João Paulo Gomes, Carly Aquino, Isabella M. Savino, Karinda Felton, Moneeb Bajwa, Nyjil Hayward, Holly Miller, Allison Naumann, Ria Allman, Neel Greer, Amary Fall, Heba H. Mostafa, Martin P. McHugh, Daniel M. Maloney, Rebecca Dewar, Juliet Kenicer, Abby Parker, Katharine Mathers, Jonathan Wild, Seb Cotton, Kate E. Templeton, George Churchwell, Philip A. Lee, Maria Pedrosa, Brenna McGruder, Sarah Schmedes, Matthew R. Plumb, Xiong Wang, Regina Bones Barcellos, Fernanda M.S. Godinho, Richard Steiner Salvato, Aimee Ceniseros, Mallery I. Breban, Nathan D. Grubaugh, Glen R. Gallagher, Chantal B.F. Vogels

## Abstract

The 2022 multi-country monkeypox (mpox) outbreak concurrent with the ongoing COVID-19 pandemic has further highlighted the need for genomic surveillance and rapid pathogen whole genome sequencing. While metagenomic sequencing approaches have been used to sequence many of the early mpox infections, these methods are resource intensive and require samples with high viral DNA concentrations. Given the atypical clinical presentation of cases associated with the outbreak and uncertainty regarding viral load across both the course of infection and anatomical body sites, there was an urgent need for a more sensitive and broadly applicable sequencing approach. Highly multiplexed amplicon-based sequencing (PrimalSeq) was initially developed for sequencing of Zika virus, and later adapted as the main sequencing approach for SARS-CoV-2. Here, we used PrimalScheme to develop a primer scheme for human monkeypox virus that can be used with many sequencing and bioinformatics pipelines implemented in public health laboratories during the COVID-19 pandemic. We sequenced clinical samples that tested presumptive positive for human monkeypox virus with amplicon-based and metagenomic sequencing approaches. We found notably higher genome coverage across the virus genome, with minimal amplicon drop-outs, in using the amplicon-based sequencing approach, particularly in higher PCR cycle threshold (lower DNA titer) samples. Further testing demonstrated that Ct value correlated with the number of sequencing reads and influenced the percent genome coverage. To maximize genome coverage when resources are limited, we recommend selecting samples with a PCR cycle threshold below 31 Ct and generating 1 million sequencing reads per sample. To support national and international public health genomic surveillance efforts, we sent out primer pool aliquots to 10 laboratories across the United States, United Kingdom, Brazil, and Portugal. These public health laboratories successfully implemented the human monkeypox virus primer scheme in various amplicon sequencing workflows and with different sample types across a range of Ct values. Thus, we show that amplicon based sequencing can provide a rapidly deployable, cost-effective, and flexible approach to pathogen whole genome sequencing in response to newly emerging pathogens. Importantly, through the implementation of our primer scheme into existing SARS-CoV-2 workflows and across a range of sample types and sequencing platforms, we further demonstrate the potential of this approach for rapid outbreak response.

## Introduction

The integration of pathogen whole genome sequencing with public health surveillance provides a powerful tool to inform outbreak control [1,2]. While the feasibility of real-time genomic surveillance was demonstrated during the 2013-2016 Ebola outbreak [3], the SARS-CoV-2 pandemic has launched a revolution in viral genomics [4]. To date, more than 14 million SARS-CoV-2 genomes have been sequenced and shared publicly [5], furthering our understanding of viral transmission and evolution. The rapid advancement in pathogen genomics in public health laboratories was facilitated in the United States through significant investments by the Centers of Disease Control and Prevention’s Office of Advanced Molecular Detection. These programs situated sequencing equipment in state and local public health laboratories, and provided practical training in laboratory and bioinformatics approaches to allow for the rapid adoption of new sequencing methods. As the COVID-19 pandemic remains ongoing, the recent spread of human monkeypox virus outside of endemic areas has provided a new target for genomic surveillance efforts [6].

Monkeypox is a zoonotic DNA virus of the *Orthopox* genus endemic to Western and Central Africa [7]. The virus consists of 3 major clades (I, IIa, and IIb), with a subgroup of clade IIb being referred to as human monkeypox virus due to direct transmission from human to human [8]. Initially rare outside endemic countries beyond imported cases, human monkeypox virus has emerged as a global threat. It was first detected in the United Kingdom on May 7, 2022, quickly spreading to other continents through travel-related infections and sometimes unknown transmission chains [9]. As of January 5, 2023, 84,075 cases of monkeypox across 103 non-endemic countries have been reported, often with atypical clinical presentations [6,10]. The lack of consistent clinical presentations, unknown transmission dynamics, and uncertainty surrounding animal reservoirs highlights the importance of establishing rapid genomic surveillance networks.

Much of the early human monkeypox virus sequencing during the 2022 global outbreak has been accomplished via a metagenomics approach [9], which employs sequencing of the total nucleic acid present in a sample. While considered the gold standard for untargeted sequencing, metagenomics incurs a high resource cost, requires experienced sequencing and bioinformatics teams, is computationally demanding, and relies on samples with high viral concentrations relative to background nucleic acids [11]. The initial metagenomic approaches included hybrid assemblies of both long and short read sequencing to allow for high confidence and polishing of the early genomes. Although this was important to establish reference genomes, it comes at the cost of being able to sequence larger numbers of specimens [12]. These characteristics of metagenomic sequencing make the approach less suitable for rapid response to large-scale pathogen outbreaks, which require low-cost, rapidly deployable solutions able to be implemented across a range of settings, sample types, and experience levels.

Highly multiplexed amplicon-based sequencing (PrimalSeq) was initially developed to generate greater coverage and depth for Zika virus genomes [13], and was later adapted as the main sequencing approach during the SARS-CoV-2 pandemic for both Illumina and Oxford Nanopore Technologies sequencing platforms [14]. Here, we developed a primer scheme for human monkeypox virus using PrimalScheme for use with amplicon-based sequencing workflows widely established during the COVID-19 pandemic. We show a consistently high percent or complete genome coverage across a range of PCR cycle-threshold (Ct) values, with different workflows and sequencing platforms. Our amplicon-based sequencing approach provides a more sensitive, lower cost, and higher throughput strategy that can be “plugged” into currently established genomic infrastructure, providing an invaluable tool for public health surveillance of human monkeypox virus.

## Results

In May 2022, a growing cluster of monkeypox cases in humans was reported outside its endemic region [6,10]. Difficulties in obtaining sufficient coverage with metagenomic sequencing approaches led us to develop a primer scheme for use with amplicon-based sequencing approaches. Given that many of the early B.1 outbreak clade genomes had low coverage, we used the closely related pre-outbreak A.1 clade genome (GenBank accession: MT903345) as a reference for the primer scheme. The primer scheme, designed using PrimalScheme [15], consists of 163 primer pairs with an amplicon length ranging between 1597 and 2497 bp (average length of 1977 bp; **Supplementary Table 1**). For the initial validation, we sequenced 10 clinical specimens with a range of PCR Ct values with both amplicon-based and metagenomic sequencing approaches at the Massachusetts State Public Health Laboratory (MASPHL). Clinical specimens ranged in Ct value from 15.0 (highest DNA concentration) to 34.6 (lowest DNA concentration). We found comparable genome coverage between amplicon and metagenomic sequencing with low Ct (<18) samples, but dramatically higher genome coverage with amplicon sequencing in higher Ct samples (>18; **Figure 1**) as compared to metagenomics sequencing. These initial findings show that amplicon-based sequencing approaches can help to improve genome coverage of human monkeypox virus genomes from clinical specimens, particularly at higher Ct values.

**Figure 1:**
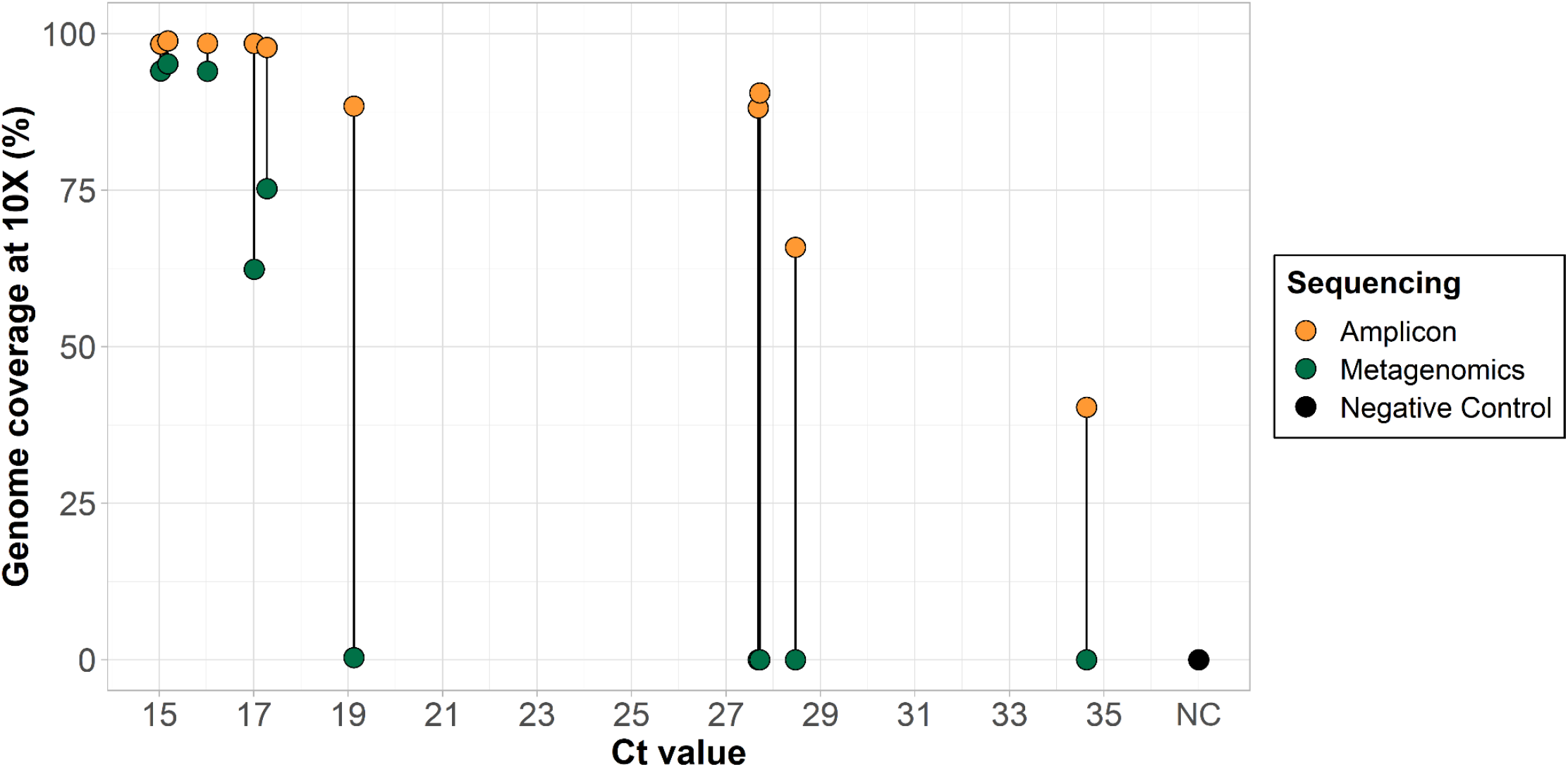
Comparison of percent genome coverage at 10X of clinical specimens sequenced with amplicon-based and metagenomic sequencing approaches. DNA was extracted from 10 clinical samples manually extracted with the QIAamp DSP DNA Blood Mini kit and PCR cycle threshold (Ct) values were determined with the non-variola *Orthopox* real-time PCR assay. Libraries were prepared with amplicon-based and metagenomic sequencing approaches and sequenced on the Illumina MiSeq (2×150 bp) with a targeted 0.5-1 million reads per library for amplicon-based sequencing and 1.5-3 million reads per library for metagenomic sequencing. A negative template control was included during library prep for each sequencing run. For amplicon-based sequencing, consensus genomes were generated at a read depth coverage of 10X and percent genome coverage as compared to the reference genome (MT903345) was determined using TheiaCoV_Illumina_PE Workflow Series on Terra.bio. For metagenomic sequencing, genomes were generated using the Broad Institute’s viral-pipelines workflows on Terra.bio using both the assemble_refbased and assemble_denovo workflows.

To further test the primer scheme, we sequenced an additional 145 clinical specimens in two independent laboratories (**Figure 2**). These samples consisted of 115 lesion swabs and 8 oropharyngeal swabs sequenced at the MASPHL, and 22 lesion swabs collected by the Connecticut Department of Public Health (CDPH) and sequenced at the Yale School of Public Health (YSPH). Our sequencing results from both laboratories confirmed that we were able to successfully sequence human monkeypox virus from lesion swabs with a range of Ct values using two different library prep kits (Illumina DNA prep (MASPHL) and Illumina COVIDSeq test (CDPH/YSPH) kits). Lesion swabs were the most common sample type used, as this sample type is recommended by the U.S. Food & Drug Administration (FDA) for clinical diagnostics [16]. However, several studies have shown that human monkeypox virus can also be detected in other sample types such as the throat or oropharyngeal swabs, saliva, feces, urine, and semen [17,18]. MASPHL had access to alternate sample types and sequenced 8 oropharyngeal swabs with or without the presence of lesions in the throat. Although the total number of samples is low, we show that near-complete genomes can be sequenced from oropharyngeal swabs, in the presence and absence of lesions (**Figure 2A**). This shows that both lesion and oropharyngeal swabs could serve as sample types for sequencing.

**Figure 2:**
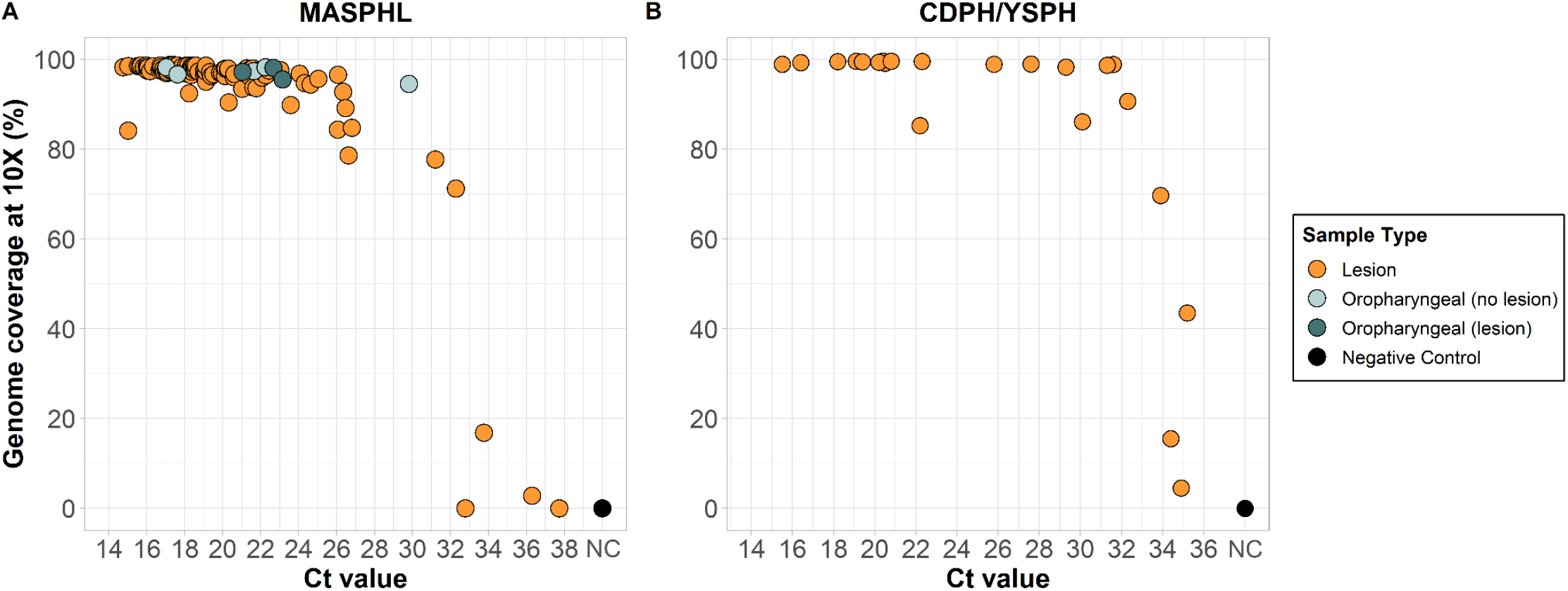
Percent genome coverage at 10X for clinical specimens sequenced with the amplicon-based sequencing approach. **A**. Clinical specimens (n=123) consisting of 115 lesion swabs, 5 oropharyngeal swabs in the absence of lesions, and 3 oropharyngeal swabs in the presence of lesions sequenced by the Massachusetts State Public Health Laboratory (MASPHL). Libraries were prepared using the Illumina DNA prep kit and sequenced on the MiSeq with 0.5-1 million reads per sample. A negative template control was included during library prep for each sequencing run. **B**. Lesion swabs (n=22) obtained from 12 individuals through the Connecticut Department of Public Health (CDPH) and sequenced by the Yale School of Public Health (YSPH). Libraries were prepared using the Illumina COVIDSeq test (RUO version) and sequenced on the NovaSeq with on average 12 million reads per sample. A negative template control was included during library prep. Bioinformatic analyses were unified between both laboratories using iVar with a minimum read depth of 10.

While both laboratories utilized the same primer scheme, there was a greater variation in percent genome coverage at lower Ct values in the MASPHL data compared to CDPH/YSPH. This difference in coverage between the two laboratories may be due to the difference in sequencing reads allotted to each sample. MASPHL sequenced on the Illumina MiSeq with 0.5-1 million reads/sample, whereas CDPH/YSPH sequenced on the Illumina NovaSeq which generated a higher output resulting in on average ∼12 million reads per sample (range: ∼0.4 to ∼20 million reads). To further investigate the optimal number of sequencing reads per sample, we randomly down-sampled the 22 clinical specimens sequenced by CDPH/YSPH to 0.5, 1, 1.5, and 2 million sequencing reads per sample. We chose data from CDPH/YSPH for downsampling because the large number of initial reads per sample allowed for a wider range of downsampling read depths than would be possible with the data from MASPHL. To better understand the threshold for sequencing, we then used a logistic function analysis to determine the PCR Ct value threshold to achieve 80% genome coverage at 10X read depth (i.e. at least 10 sequencing reads aligned to a genome position) for each level of downsampling. We found that down-sampling the total number of sequencing reads from 2 million to 0.5 million decreased the 80% coverage Ct threshold by 1 Ct (from 32.3 Ct to 31.3 Ct), and resulted in an average 7.2% (range: 0-26.1%) decrease in percent genome coverage at 10X (**Figure 3**). To achieve an overall high genome coverage of more than 80% at 10X read depth, we recommend generating at least 1 million sequencing reads per sample, if resources allow. Further increasing the number of sequencing reads to 2 million helps to generate higher coverage for samples with relatively high Ct values >31, but only small differences in coverage at 10X read depth of on average 2.1% were observed with samples that had Ct values <31. Thus, we recommend selecting samples for sequencing with a Ct value below 31 and generating at least 1 million reads per sample, to maximize genome coverage, particularly when resources are limited.

**Figure 3.**
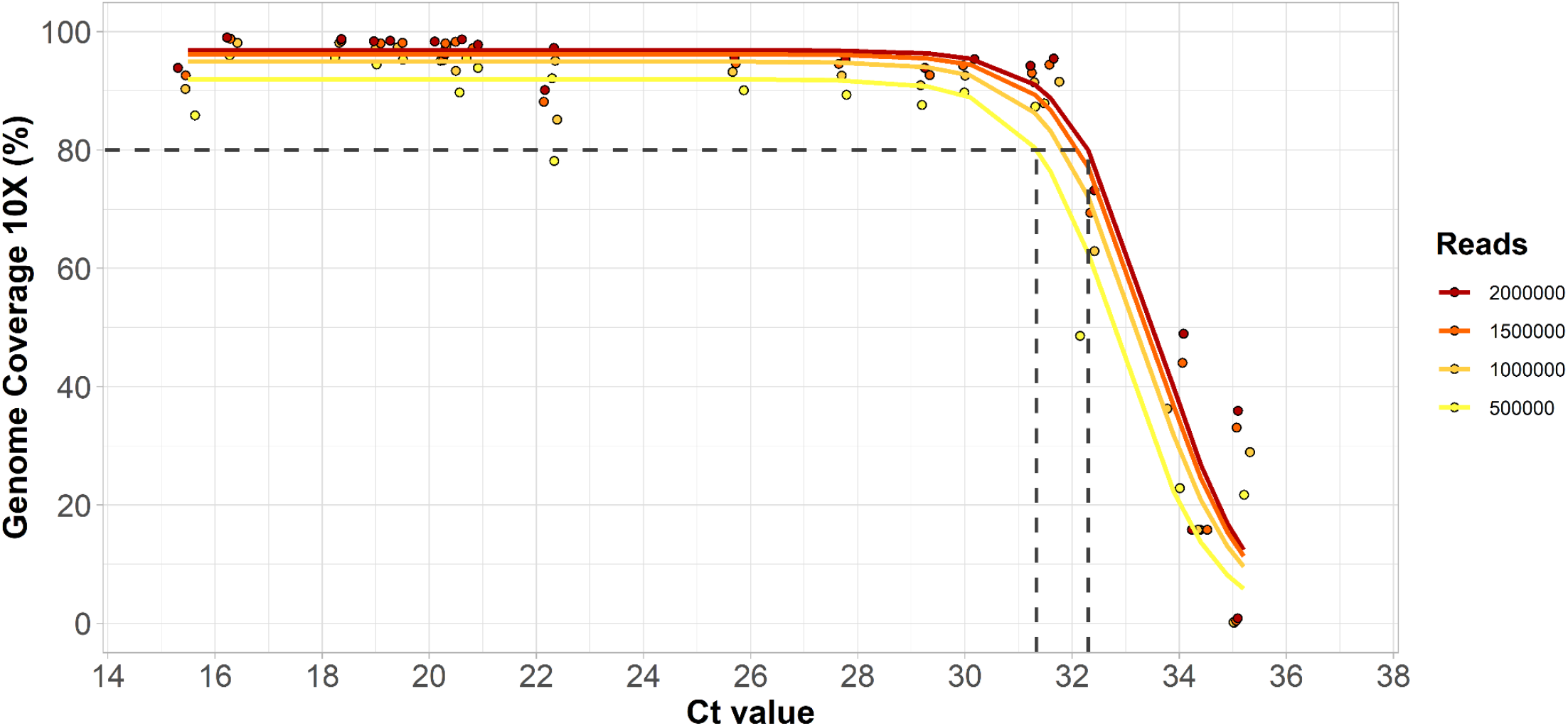
Percent genome coverage at 10X mapped read depth for 22 clinical specimens after randomly down-sampling to a specific number of sequencing reads. To further investigate the combined effects of Ct value and number of sequencing reads per sample, we randomly down-sampled the CDPH/YSPH data to 2, 1.5, 1, and 0.5 million reads per sample. We used a logistic function analysis to plot the fitted lines indicating the decrease in percent genome coverage with higher Ct values.

To investigate the differences in coverage depth for the randomly down-sampled data, we determined the depth of coverage at each nucleotide position across the genome. This analysis again confirmed that increasing Ct values and decreasing sequencing reads per sample result in more regions of the genomes with coverage <10X depth (**Figure 4**). Moreover, it showed variation in depth of coverage across the genome. To further investigate which specific amplicons consistently had low coverage, we determined the depth of coverage for non-overlapping regions of amplicons in our dataset that was randomly down-sampled to 1 million reads/sample and for samples with a Ct <31. By excluding genomic regions covered by overlapping amplicons, we found that amplicons 75 and 118 consistently had a depth of coverage <10X across all 15 samples included in the analysis. Additionally, the mean depth of coverage for amplicons 11, 26, 28, 56, 59, 60, 74, and 96 was also below 10X. To understand the cause for the lower coverage, we investigated whether there were any mismatches in the primers for these amplicons. This revealed a single mismatch with the MPXV_11_RIGHT primer at the 3’ end, whereas no mismatches were present in any of the other primers. This suggests that differences in PCR efficiency may explain the lower coverage across these amplicons. Further optimization would be required to improve uniformity in PCR efficiency across the primer pairs in the current primer scheme. This may also help to reduce the number of sequencing reads required to reach a minimum coverage of 10X across the entire genome.

**Figure 4:**
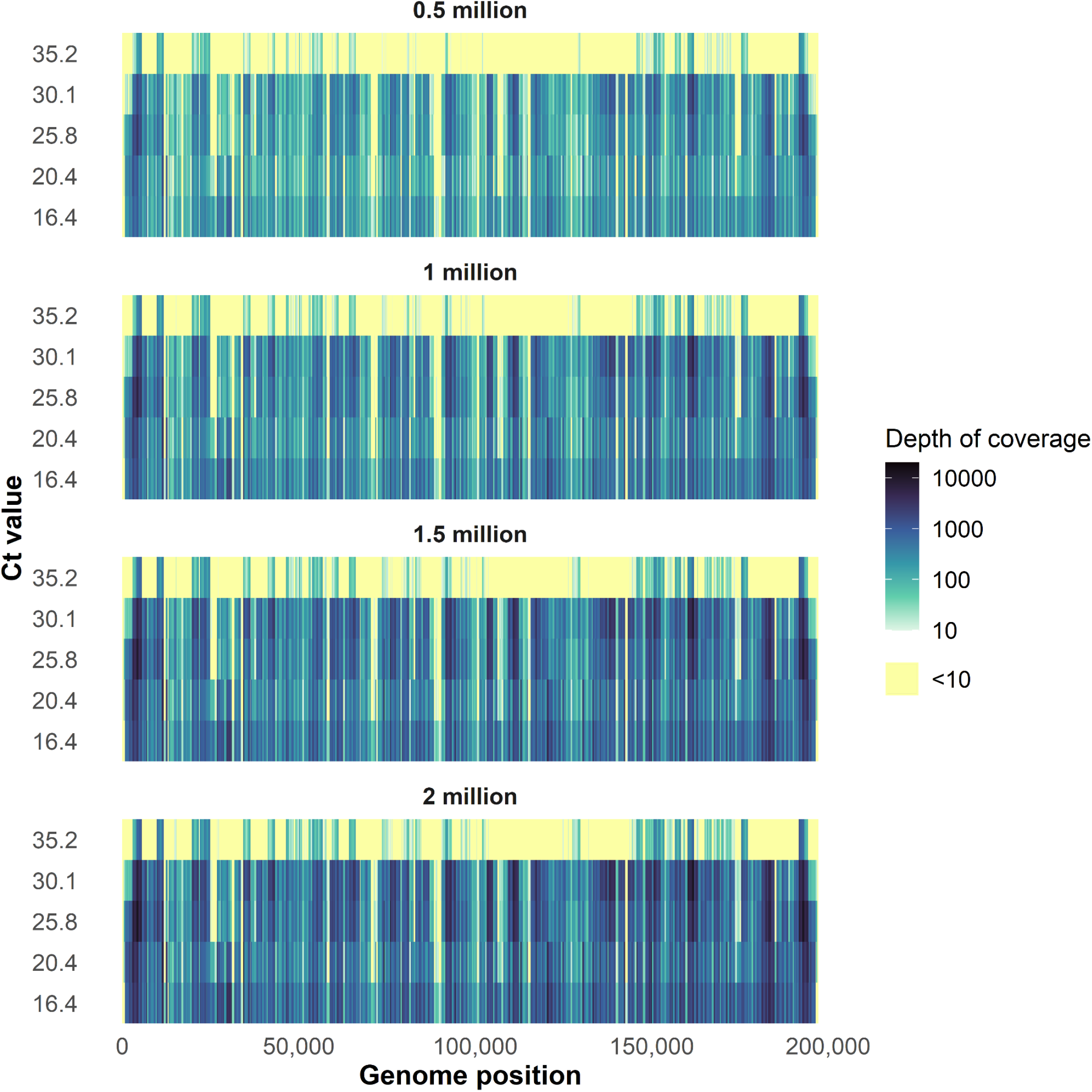
Depth of coverage by genome position for samples representing a range of Ct values and randomly down-sampled to different numbers of sequencing reads. We determined the depth of coverage at each nucleotide position for selected samples that represented a range in Ct values from 16.4-35.2, and for which the number of raw sequencing reads was randomly down-sampled to 2, 1.5, 1, and 0.5 million sequencing reads. Each row represents a single specimen, ranked by Ct value from high (low DNA titer) to low (high DNA titer). Highlighted in yellow are positions of the genome with a depth of coverage below 10X.

To support global outbreak response efforts, we immediately made our primer scheme and protocols publicly available following validation [19], and shipped primer pool aliquots to 10 additional public health laboratories across the United States and internationally (**Figure 5**). Each laboratory “plugged” the human monkeypox virus primers into their established SARS-CoV-2 amplicon-based sequencing workflow and sequenced their samples on Illumina or Oxford Nanopore Technologies platforms. We unified the data analysis for all laboratories by running the same bioinformatics pipeline (iVar) to generate consensus genomes and to determine percent genome coverage at 10X read depth. Despite variation between workflows and sequencing platforms, we found that amplicon-based sequencing resulted in relatively consistent high genome coverage (>80%), with decreasing coverage at higher Ct values (**Figure 6**). By using a logistic function analysis, we determined the PCR Ct value threshold at which each laboratory was able to achieve 80% genome coverage at 10X read depth, which ranged between Ct 24.7 to 31.24 **(Figure 6A-C, I)**. For some laboratories we were not able to determine this threshold, because (1) the range in Ct values was too narrow to see the decrease at higher Ct values (**Figure 6D-G**), or (2) there was high variation in the coverage at lower Ct values (**Figure 6H, J**). The reason why some samples with low Ct values failed to achieve high genome coverage is unknown, but this was likely due to technical error during library preparation or sequencing, and not due to the primer scheme itself as such patterns would have been consistent across all the laboratories. Most samples were sequenced on Illumina platforms, as this was the most commonly used sequencing platform in the laboratories that requested primers. Based on our previous experience with amplicon-based sequencing of viruses such as Zika and SARS-CoV-2, we expected that the primer scheme could also be used with Oxford Nanopore Technologies workflows. The NHS Lothian laboratory had the ability to prepare libraries with both Illumina and Oxford Nanopore technologies workflows and sequenced on the Illumina MiSeq and Oxford Nanopore Technologies GridION. This pairwise comparison showed that genome coverage at 10X read depth was comparable between both workflows (**Figure 6I**). By comparing data generated by independent public health laboratories, we show that the human monkeypox virus primer scheme can be successfully implemented in previously established Illumina and Oxford Nanopore Technologies sequencing workflows under varying conditions.

**Figure 5.**
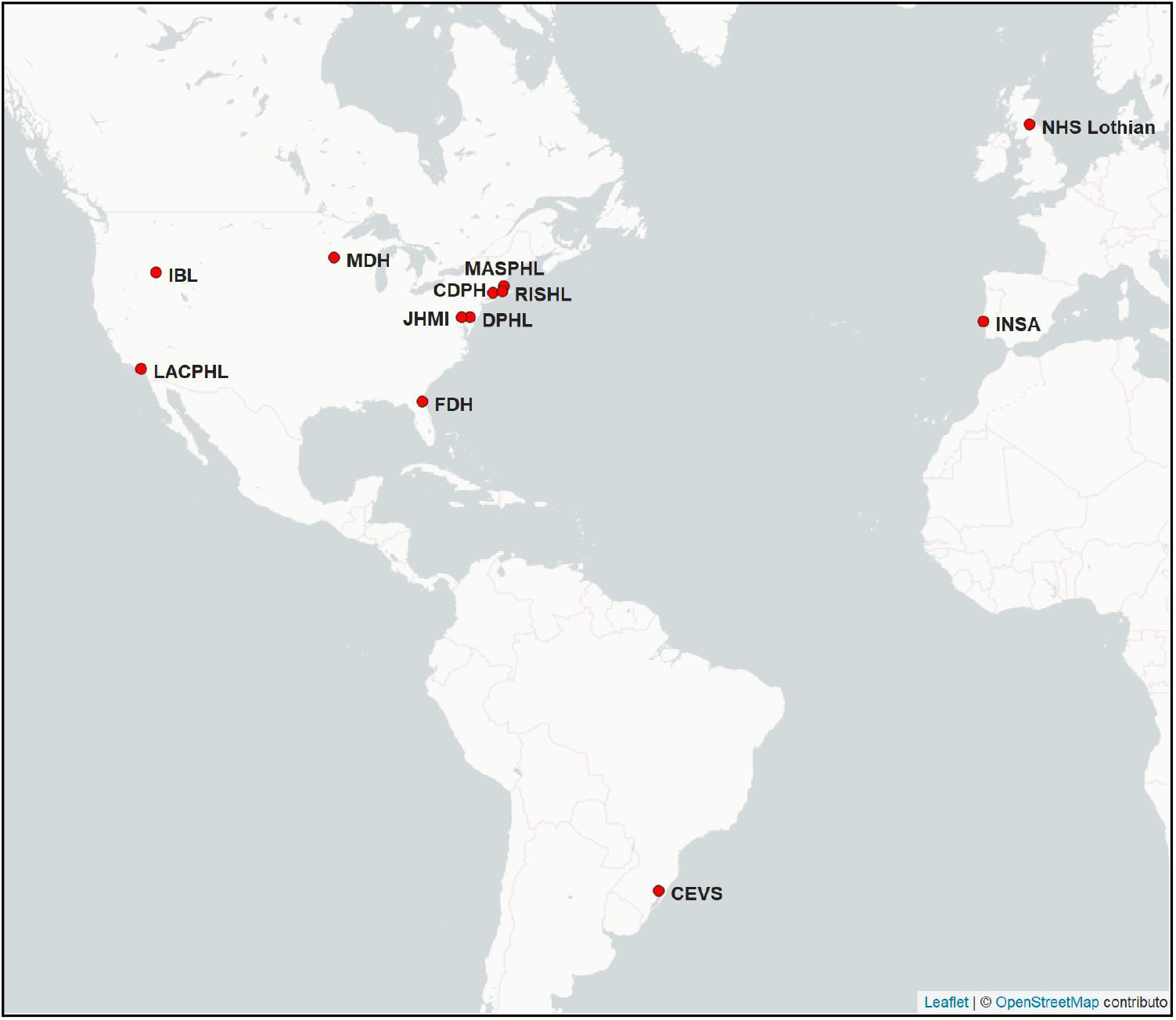
Geographical distribution of public health laboratories that implemented the human monkeypox virus primer scheme with their established amplicon-based sequencing workflows. Public health laboratories contributing data to this study include: Connecticut Department of Public Health (CDPH), Centro Estadual de Vigilância em Saúde (CEVS), Delaware Public Health Lab (DPHL), Florida Department of Health (FDH), Idaho Bureau of Laboratories (IBL), Johns Hopkins Medical Institutions (JHMI), Los Angeles County Public Health Lab (LACPHL), Massachusetts State Public Health Laboratory (MASPHL), Minnesota Department of Health (MDH), National Health Service Lothian (NHS Lothian), National Institute of Health Dr. Ricardo Jorge (INSA), and Rhode Island State Health Laboratory (RISHL).

**Figure 6:**
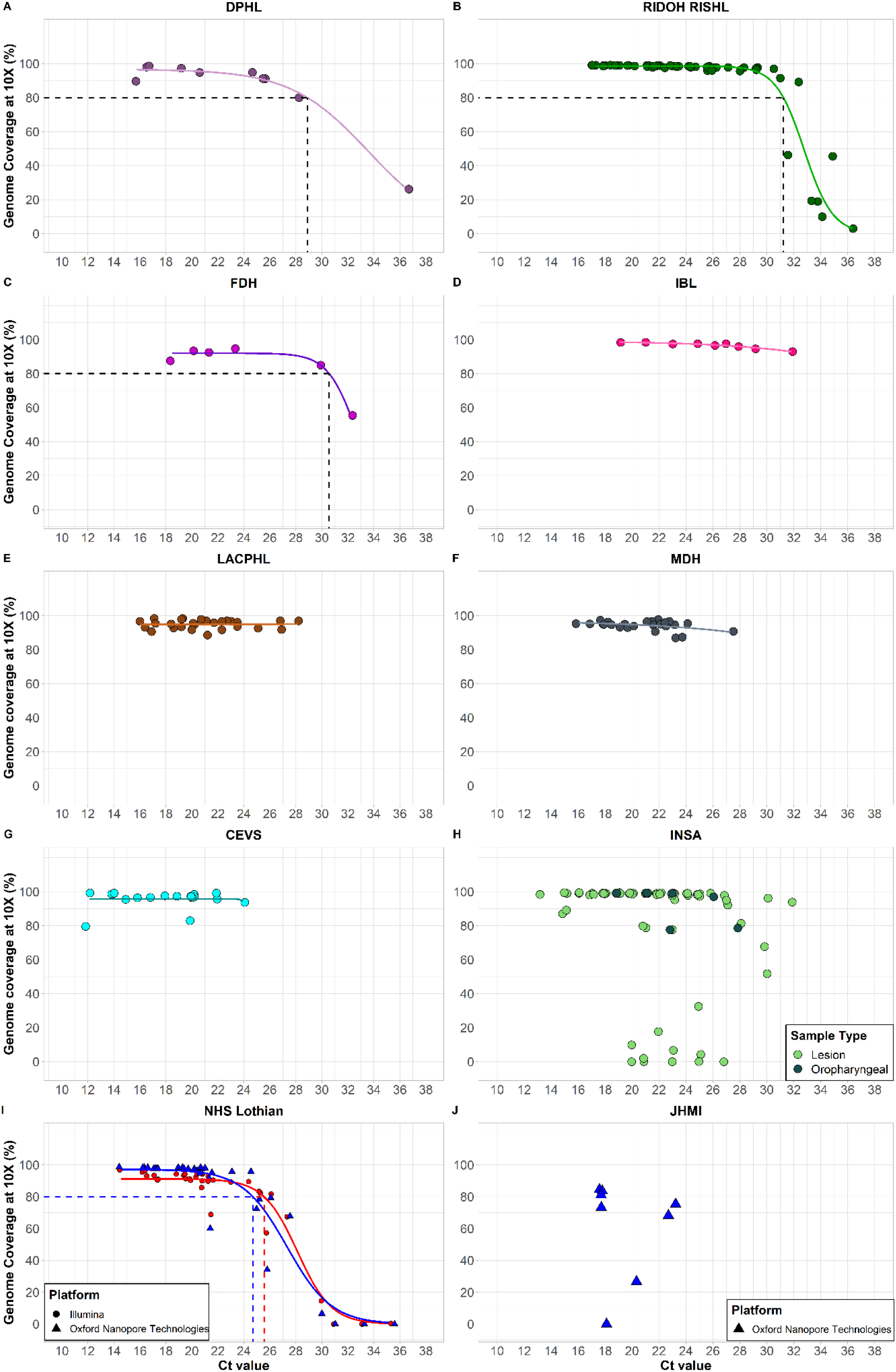
Percent genome coverage at 10X read depth for clinical specimens sequenced with the amplicon-based sequencing approach. **A**. Lesion swabs (n=10) sequenced by the DPHL. Data are fitted with a logistic function and the dashed line corresponds to 80% genome coverage at a threshold of Ct 28.4. **B**. Dry vesicle swabs (n=56) from various anatomical sites and sequenced by the RIDOH RISHL. Data are fitted with a logistic function and the dashed line corresponds to 80% genome coverage at a threshold of Ct 31.2. **C**. Lesion swabs (n=6) sequenced by the FDH. Data are fitted with a logistic function and the dashed line corresponds to 80% genome coverage at a threshold of Ct 30.6. **D**. Lesion swabs (n=9) sequenced by IBL. Data are fitted with a logistic function. **E**. Lesion swabs (n=27) sequenced by the LACPHL. Data are fitted with a logistic function. **F**. Lesion swabs (n=25) from various anatomical sites and sequenced by the MDH. Data are fitted with a logistic function. **G**. Clinical specimens (n=19) consisting of lesion swabs and crusts of healing lesions sequenced by the CEVS. Data are fitted with a logistic function. **H**. Clinical specimens (n=78) consisting of lesion swabs from various anatomical sites as well as oropharyngeal swabs sequenced by INSA. **I**. Vesicle swabs (n=34) from various anatomical sites tested in parallel on Illumina and Oxford Nanopore Technology (ONT) sequencing platforms by the NHS. Data from both sequencing platforms are fitted with logistic function and the dashed lines correspond to 80% genome coverage at a threshold of Ct 25.0 on the Illumina platform and Ct 24.4 on the ONT platform. **J**. Lesion swabs (n=8) sequenced on the ONT platform by JHMI.

## Discussion

We developed an amplicon-based sequencing approach for human monkeypox virus to provide a more sensitive, lower cost, and higher throughput alternative to metagenomic sequencing. We used PrimalScheme to develop primers for human monkeypox virus based on a pre-outbreak A.1 lineage genome (GenBank accession: MT903345) and tested the primer scheme with clinical specimens in two independent laboratories. After initial validation, we shared primer pool aliquots with 10 additional public health laboratories, who successfully implemented the scheme in their existing amplicon-based sequencing workflows. Our findings showed greater breadth and depth of genome coverage when using the amplicon-based sequencing approach as compared to metagenomics, particularly for specimens with lower DNA concentrations. We identified Ct value and number of sequencing reads as two factors that influence percent genome coverage. Based on our findings we made the following recommendations for amplicon-based sequencing of human monkeypox virus:

1. Utilize existing amplicon-based SARS-CoV-2 sequencing infrastructure
2. Prioritize samples with a Ct value <31, if resources are limited
3. Generate at least 1 million sequencing reads per sample, if resources allow

Importantly, our human monkeypox virus primer scheme can be used with currently implemented SARS-CoV-2 sequencing workflows and Illumina or Oxford Nanopore Technologies sequencing platforms. This will enable laboratories that are currently using amplicon-based sequencing approaches for SARS-CoV-2 to expand their portfolio by including human monkeypox virus, with minimal change to their overall sequencing and bioinformatics workflow.

The comparatively steep drop-off in genome coverage with higher PCR Ct samples seen with metagenomics reinforces a limitation of this sequencing approach found in other studies that used clinical metagenomics for pathogen sequencing [20]. This lack of sensitivity presents a potential challenge to genomic surveillance, as human monkeypox virus DNA concentrations fluctuate throughout the course of infection and across specimen types [21]. We show that high genome coverage can be achieved with different workflows, sequencing platforms, and sample types such as lesion and oropharyngeal swabs. The higher sensitivity seen in amplicon-based sequencing allows for sequencing of a larger variety of sample types across a wider range of Ct values compared to metagenomic sequencing.

Given the large number (163) of amplicons needed to span the entire human monkeypox virus genome, there was an increased potential for amplicon drop-outs and a resulting reduction in genome coverage. Despite being an inherent risk of amplicon sequencing, here we identified only two amplicons (75 and 118) with consistent low coverage <10X, and an additional 8 amplicons with overall lower coverage. This lower coverage is likely the result of lower amplification efficiency, and may be improved by further optimization to achieve high PCR efficiency across all the primer sets. As a double-stranded DNA virus with genetic proof-reading mechanisms, the monkeypox virus has a comparatively slower evolutionary rate than single-stranded RNA viruses [22]. Less genetic diversity over time translates to fewer differences between the reference genome used to generate the primer scheme and genomes that are associated with the 2022 outbreak, decreasing the risk for amplicon drop-out. As a result, the primer scheme may not have to be updated as frequently as, for example, the primer scheme used to sequence SARS-CoV-2 [14].

Monkeypox virus exhibits high genetic diversity highlighted by the presence of deep phylogenetic branches between different clades [8]. Our scheme is designed to target the viruses associated with the global outbreak (clade IIb), which is characterized by high human-to-human transmission, to facilitate genomic surveillance. Although the performance of our scheme has not been verified for sequencing of viruses belonging to other clades (I and IIa) typically found in the endemic regions, we anticipate a lower genome coverage compared to strains associated with the current outbreak due to drop-out of primers found in regions with a high genetic divergence between the clades. Therefore, the development of additional schemes targeting other endemic clades is needed in the future to cover all monkeypox virus clades. The availability of such schemes will facilitate their broad applicability in genomic surveillance of the strains associated with the global outbreak and those locally circulating in endemic regions, such as in Central and West Africa. One of the main advantages of our scheme is its “plug-and-play” design, which will allow the reuse of pre-existing trained personnel and infrastructure developed for the SARS-CoV-2 genomic surveillance. This will ensure rapid and seamless implementation of the current scheme and additional versions targeting other clades, especially those commonly found in endemic countries. Ultimately, these efforts may increase the availability of genomic data from endemic regions.

There were several limitations in this study. First, the large number of primers included in the scheme resulted in differences in PCR efficiency between amplicons. By down-sampling the sequencing data, we show that a higher number of sequencing reads can overcome these challenges. Further optimization by designing primers targeting other positions and changing primer concentrations may help to reduce the number of raw sequencing reads needed to reach at least 10X coverage depth across the genome. Second, although results between laboratories were consistent, we observed some variability in the relation between Ct value and genome coverage. Differences in the sample types, qPCR assays, sequencing workflows, and thermocycler calibration can likely explain this variability. Third, there were some challenges with the more variable ends of the genome that contain inverted terminal repeats. This resulted in multiple binding sites for primers spanning these regions. In addition, this can create challenges when mapping reads to the inverted terminal repeats for both Illumina and Oxford Nanopore Technologies data, which may result in the misalignment of reads. This warrants careful interpretation of sequencing results in these genome regions. Lastly, Orthopox viruses are known to have genomic rearrangements (translocations, duplications, and inversions) especially in the inverted terminal repeats [23], which may not be well identified by amplicon-based sequencing. Although we recently identified a 600 bp deletion in the inverted terminal repeats that affects commonly used real-time PCR assays [24], amplicon-based sequencing may not be able to identify other genomic rearrangements that may have epidemiological or clinical significance. We recommend periodic long read metagenomic sequencing to supplement large-scale amplicon-based sequencing as an additional strategy to improve genomic surveillance of human monkeypox virus.

Through this study, we have shown that amplicon-based sequencing can increase the sensitivity, breadth, and depth of human monkeypox virus genome coverage with low DNA concentration specimens. By being able to “plug” the human monkeypox virus primer scheme into existing sequencing and bioinformatics infrastructure, our approach has helped public health laboratories worldwide to quickly adapt their existing workflows in response to the global mpox outbreak.

## Methods

### Ethics statement

As part of this study, we sequenced remnant clinical specimens that tested presumptive positive for monkeypox virus. Ethical oversight for each institution is indicated in **Table 1**. All data were de-identified prior to sharing and sample codes as included in the manuscript are not known outside the research groups.

**Table 1:**
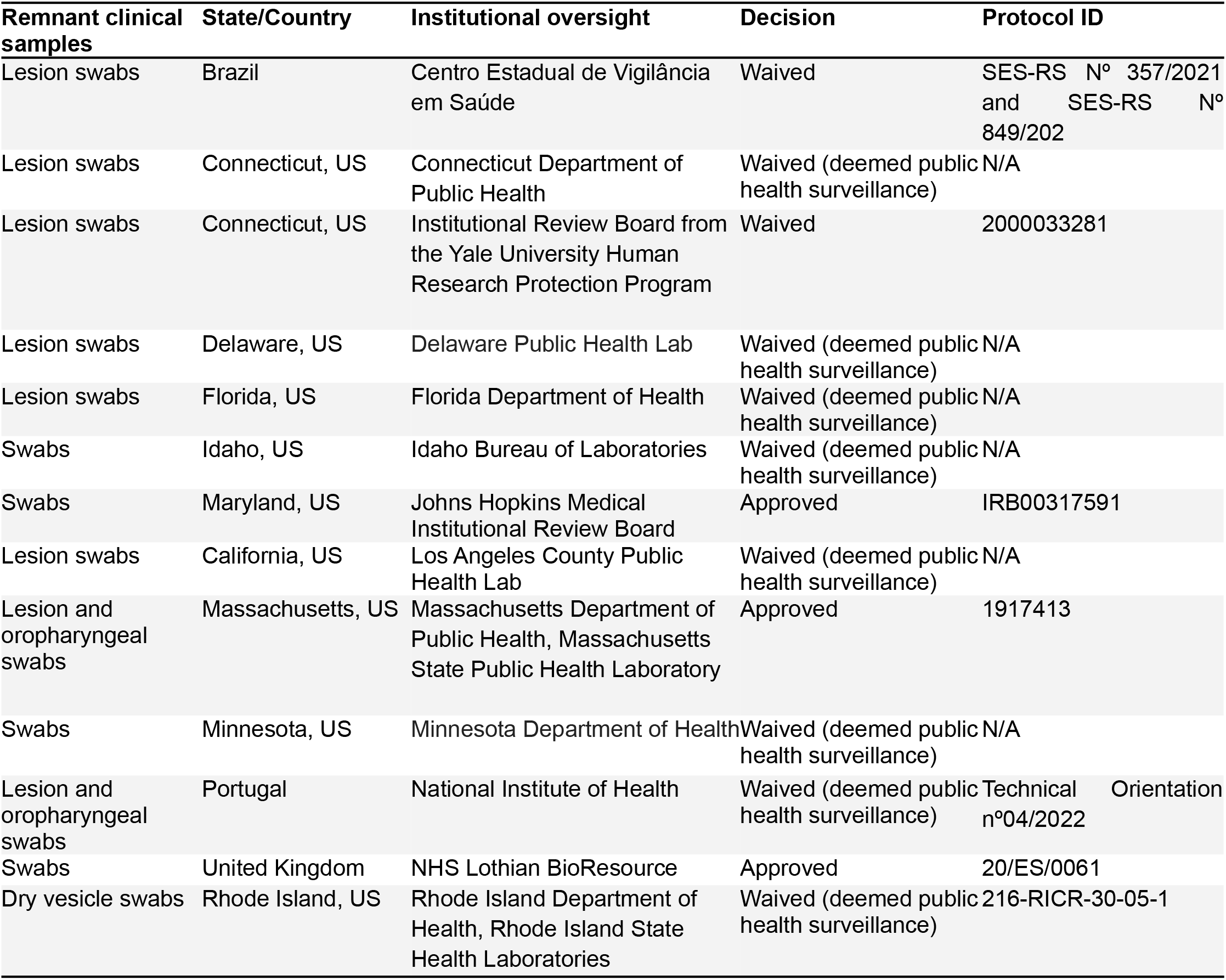
Ethical oversight.

### Primer scheme design

The human monkeypox primer scheme (v1) was designed with PrimalScheme [15] using a pre-outbreak clade IIb reference genome (GenBank accession: MT903345) belonging to the A.1 lineage, following the newly proposed monkeypox virus naming system [8]. The primer scheme comprises a total of 163 primer pairs with an amplicon length ranging between 1597 and 2497 bp (average length of 1977 bp; **Supplementary Table 1**).

### Clinical specimens for validation

Validation of the primer scheme was done by MASPH, CDPH, and YSPH. At CDPH, a total of 22 clinical specimens consisting of swabs from lesions of 12 individuals were tested. DNA was extracted from clinical specimens via the Roche MagNA pure 96 kit or manual extraction and tested with the non-variola *Orthopox* real-time PCR assay on the Bio-Rad CFX96 instrument [25].

At MASPHL, a total of 133 clinical specimens consisting of both lesion swabs and oropharyngeal swabs with and without detection of lesions in the throat were tested. DNA was extracted from clinical specimens using the QIAamp DSP DNA Blood Mini kit and tested with the non-variola *Orthopox* real-time PCR assay on the ABI 7500 Fast Real Time PCR instrument [25].

### Metagenomic sequencing

Initial validation was done by sequencing 10 clinical specimens with both metagenomic and amplicon-based sequencing approaches at MASPHL. For metagenomic sequencing, samples were first quantified using the Qubit 1x dsDNA High Sensitivity kit to determine concentration. Initial loading volume was calculated for each sample to fall in the recommended range of 100 to 500 ng. Each sample was then run in duplicate through the Illumina DNA Prep kit for library preparation following manufacturer’s protocol. After Flex Amplification PCR, the libraries were cleaned using the protocol option for standard DNA input. Post-library purification, each sample library was run on the Tapestation D1000 to determine the average peak size. Samples were also again quantified using the Qubit dsDNA High Sensitivity kit to best normalize the samples when pooling the libraries. Samples were then pooled, denatured, and diluted to 10 pM before being spiked with 5% PhiX. The diluted pooled libraries were then loaded and run on an Illumina v.2 300 cycle MiSeq cartridge, with a target of 1.5-3 million reads per library. Genomes were generated using the Broad Institute’s viral-pipelines workflows on Terra.bio using both the assemble_refbased and assemble_denovo workflows. The assemble_refbased workflow aligned reads against the MA001 (ON563414.3) genome for consensus generation. The assemble_denovo workflow scaffolded de novo SPAdes contigs against both the MA001 (ON563414.3) and RefSeq (NC_063383.1) references, followed by read based polishing.

### Amplicon-based sequencing

At MASPHL, libraries were prepared for sequencing using the Illumina DNA prep kit [19]. Initial library clean-up in the Illumina DNA prep protocol was done by following recommendations for standard DNA input, but later optimization showed improved coverage when following the recommendations for small PCR amplicon input. A negative template control was included during library prep for each sequencing run. Pooled libraries were sequenced on the Illumina MiSeq (paired-end 150), with a target of 0.5-1 million reads per library. For the initial 10 sequenced samples, primers were trimmed and consensus genomes were generated at a minimum depth of coverage of 10X via the TheiaCoV_Illumina_PE Workflow Series on Terra.bio. TheiaCoV_Illumina_PE was originally written to enable genomic characterization of SARS-CoV-2 specimens from Illumina paired-end amplicon read data. Modifications to TheiaCoV_Illumina_PE were made to to support MPXV genomic characterization; these modifications accomodated the use of a MPXV reference sequence and primer scheme for consensus genome assembly. These updates were made available in the TheiaCov v2.2.0 release [26].

At YSPH, libraries were prepared for sequencing using the Illumina COVIDSeq test (RUO version) [19]. A negative template control was included during library prep for each sequencing run. Pooled libraries were sequenced on the Illumina NovaSeq (paired-end 150), with on average 12 million reads per library (range: ∼0.4 to ∼20 million reads per library).

### Data analysis

Bioinformatic analyses were done by YSPH to unify the data analysis. BAM files containing mapped reads were shared, and mapped reads were extracted using bamToFastq or “BEDtools bamtofastq” (version 2.30.0) [27] into FASTQ files before downstream analysis. To start the analysis, we re-mapped the reads to the human monkeypox reference genome (GenBank accession: MT903345) using BWA-MEM (version 0.7.17-r1188) [28]. The generated BAM mapping files were sorted using SAMtools (version 1.6) [29] and then used as input to iVar (version 1.3.1) [30] to trim primer sequences from reads. The trimmed BAM files were then used to generate consensus sequences using iVar with a minimum read depth of 10. We also calculated the per-base read coverage depth using genomeCoverageBed or “BEDTools genomecov” (version 2.30.0) [27]. To calculate the percent genome coverage for each sample, we generated a pairwise whole-genome sequence alignment of each sample against the human monkeypox virus reference genome (GenBank accession: MT903345) using Nextalign (version 1.4.1) [31]. We then calculated the percentage of alignment positions, excluding ambiguous nucleotides and deletions, using BioPython (version 1.7.9) [32].

To investigate the effect of the number of sequenced reads on the percent genome coverage, we generated randomly down-sampled sequence reads for the CDPH/YSPH samples. We used the CDPH/YSPH samples because they were sequenced at sufficiently high depth to allow down-sampling at different total read counts, namely 0.5, 1, 1.5, and 2 million reads per sample. We first generated interleaved sequencing read files from the paired-end sequencing reads using seqfu (version 1.14.0) [33] and then randomly down-sampled the reads at the specified total read count thresholds using “seqtk sample” (version 1.3-r106) [34]. Similarly, we generated the consensus whole-genome sequences and calculated the percentage genome coverage as described in the previous paragraph.

All further data analysis and plotting were performed using R statistical software v4.2.0 [35] using the ggplot2 v3.3.6 [36], dplyr v1.0.9 [37], tidyr v1.2.0 [38], and cowplot v1.1.1 [39] packages. We subsetted each dataset to a coverage depth of 10X and fitted a logistic function specified as follows: y=a / (1 + exp(-b * (x - c))), where a, b, and c are parameters and ‘x’ is the PCR Ct value and ‘y’ is the percentage genome coverage or completeness. We used the ‘curve_fit’ function in the numpy Python package to fit the model [40]. To interpolate the Ct value corresponding to an 80% threshold Ct value, we rearranged the logistic equation to estimate ‘x’ given the estimated parameters a, b, and c, and genome coverage or ‘y’ of 80% at each read coverage depth.

### Public health response

After initial validation, we made a detailed protocol publicly available and shared pooled primer aliquots with laboratories across the United States and internationally to support their public health genomic efforts [19]. Each laboratory sequenced samples by adapting their SARS-CoV-2 amplicon sequencing workflows, performed their own bioinformatics, and submitted sequencing data and consensus genomes to public repositories such as NCBI or GISAID (**Supplementary Table 2**). By sharing de-identified sequencing data (e.g. coded human-depleted raw reads or BAM files containing mapped reads) and metadata (e.g. sample codes, sample types, and Ct values), we ran the same bioinformatics pipeline on all data generated on Illumina platforms to determine breadth and depth of coverage for each genome under standardized conditions, as described above.

After initial dissemination of primer aliquots, Integrated DNA Technologies created pre-pooled primer aliquots, which eliminates the need to manually pool individually ordered primers. These pools were validated by YSPH and revealed similar coverage as compared to initial results with manually pooled primers (**Supplementary Figure 1**).

#### Centro Estadual de Vigilância em Saúde (CEVS)

A total of 19 clinical specimens consisting of lesion swabs and the crusts of healing lesions were tested. DNA was extracted from clinical specimens using the Invitrogen PureLink Viral RNA/DNA Mini kit and tested with a clade-specific PCR assay on the Bio-Rad CFX opus 96 instrument. Libraries were prepared for sequencing using the Illumina DNA prep kit. Pooled libraries were sequenced on the Illumina MiSeq v3 (paired-end 150), with 2 million reads per library.

#### Delaware Public Health Lab (DPHL)

A total of 10 clinical specimens consisting of dry swabs from lesion sites were tested. DNA was manually extracted from clinical specimens using the QIAGEN QIAamp DSP DNA Blood Mini Kit. Amplification was achieved by using the Perfecta Multiplex qPCR SuperMix Low Rox PCR assay in conjunction with CDC issued Non-variola Orthopoxvirus Real-Time PCR Primer and Probe Set on the Applied Biosystems 7500 Fast Real-Time PCR instrument. Libraries were prepared for sequencing using the Illumina DNA prep kit. Pooled libraries were sequenced on the Illumina Miseq (paired-end 150), with about 1 million to 2.5 million reads per library.

#### Florida Department of Health (FDH)

A total of 6 clinical lesion swab specimens were tested. DNA was extracted from clinical specimens using the Qiagen QIAamp DSP DNA Blood Mini Kit and tested with the non-variola *Orthopox* real-time PCR assay on the BioRad T100 instrument [25]. Libraries were prepared for sequencing using the Illumina Nextera v2 kit. Pooled libraries were sequenced on the Illumina iSeq 100 v2 (paired-end 150), with 1-2.4 million reads per library.

#### Idaho Bureau of Laboratories (IBL)

A total of 9 lesion swab clinical specimens were tested. DNA was extracted from clinical specimens using the QIAGEN QIAamp DSP DNA Blood Mini kit and tested with Perfecta Multiplex qPCR SuperMix Low Rox PCR assay in conjunction with the CDC FDA-approved Non-variola Orthopoxvirus (VAC1) assay on the Applied Biosystems® 7500 Fast Dx Real-Time PCR instrument. Libraries were prepared for sequencing using the Illumina DNA prep kit. Pooled libraries were sequenced on the Illumina MiSeq, with 0.8-1.2 million reads per library.

#### Johns Hopkins Medical Institutions (JHMI)

A total of 8 clinical lesion swab specimens were tested. DNA was extracted from clinical specimens using the bioMérieux easyMag and tested with a LDT PCR assay that adopted the primer and probe sequences of the non-variola orthopoxvirus, modified from Li, et al [41]. Total reaction volume for the real-time PCR was 20 μL (5 μL of template and 15 μL master mix). The master mix contained 5 μL TaqPath 1-Step RT-qPCR Master Mix (Applied Biosystems, A15299, Waltham, MA), 4 μL water, and 1 μL of each primer (10 nm) and the probe (5 nm) in addition to 1 μL of each primer (10 nm) and the probe (5 nm) for the RNAseP internal control target. Real-time PCR was performed using Prism 7500 Detection System (Applied Biosystems) and the following cycling conditions: 1 cycle at 95.0°C for 2 minutes and 40 cycles at 95.0°C for 3 seconds and 60.0°C for 31 seconds. Libraries were prepared for sequencing using NEBNext ARTIC reagents for SARS-CoV-2 sequencing. Pooled libraries were sequenced on the Oxford Nanopore Technologies GridIon. Primers were trimmed and consensus genomes were generated at a minimum depth of coverage of 10X using the ARTIC bioinformatics pipeline [42].

#### Los Angeles County Public Health Lab (LACPHL)

A total of 27 remnant lesion swab specimens were tested. DNA was extracted using the Qiagen EZ1&2 DNA Tissue Kit from specimens that were previously tested with the non-variola *Orthopoxvirus* real-time PCR assay on the ABI 7500 FastDx instrument following Centers for Disease Control and Prevention Laboratory Response Network protocols [25]. Libraries were prepared for sequencing using the Illumina DNA prep kit. Pooled libraries were sequenced on the Illumina MiSeq v2 (paired-end 150), with 0.5-1 million reads per library.

#### Minnesota Department of Health (MDH)

A total of 25 clinical specimens consisting of lesion swabs from various anatomical sites were tested. DNA was extracted from clinical specimens using the QIAamp DSP DNA Blood Mini kit and tested with a non-variola *Orthopox* real-time PCR assay on the ABI 7500 Fast Real Time PCR instrument [25]. Libraries were prepared for sequencing using the Illumina DNA prep kit. Pooled libraries were sequenced on the Illumina MiSeq v2 (paired-end 250), with target of 500,000 reads per library. Primers were trimmed and consensus genomes were generated at a minimum depth of coverage of 10X using nf-core/viralrecon.

#### National Institute of Health Dr. Ricardo Jorge (INSA)

A total of 78 clinical specimens consisting of lesion swabs from various anatomical sites and oropharyngeal swabs were tested. DNA was extracted from clinical specimens using the MagMAX Viral/Pathogen Nucleic Acid Isolation kit and tested with a real-time PCR assay on the CFX Opus real-time PCR system [43–45]. Libraries were prepared for sequencing using the Illumina Nextera XT library prep kit. Pooled libraries were sequenced on the Illumina NextSeq 550, with 2 million reads per library.

#### National Health Service Lothian (NHS Lothian)

A total of 34 clinical specimens consisting of vesicle swabs and swabs from various anatomical sites were tested. DNA was extracted from clinical specimens using the Biomerieux Nuclisens EasyMag kit and tested with the clade-specific real-time PCR assay on the Applied Biosystems 7500 Fast Real-Time PCR instrument [46]. Illumina libraries were prepared for sequencing using the Illumina COVIDSeq test (RUO version). Pooled libraries were sequenced on the Illumina MiSeq - Micro v2 reagent kit, with 400,000 reads per library. Primers were trimmed and consensus genomes were generated at a minimum depth of coverage of 5X using the Public Health Wales Nextflow nCoV-2019 pipeline which utilizes iVar [47]. Whole genome PCR amplicons were also used to prepare Nanopore libraries using the Artic LoCost method [48], substituting Blunt TA Ligase with NEBNext Ultra II Ligation Mastermix for barcode ligation. Libraries were pooled on a single R9.4.1 flowcell and sequenced with a GridION (Oxford Nanopore Technologies) running live High Accuracy basecalling in MinKnow v21.11.6, aiming for 100,000 reads per library. Consensus genomes were generated at a minimum depth of coverage of 20x with the Artic field bioinformatics pipeline v1.2.1 and variants called with Nanopolish [47].

#### Rhode Island Department of Health/ Rhode Island State Health Laboratory (RIDOH RISHL)

A total of 56 clinical specimens consisting of dry vesicle swabs from various anatomical sites were tested. DNA was extracted from clinical specimens using the Qiagen QIAmp DSP Blood mini kit and tested with the non-variola *Orthopox* real-time PCR assay on the Applied biosystems 7500 instrument. Libraries were prepared for sequencing using the Illumina DNA prep kit. Pooled libraries were sequenced on the Illumina MiSeq (paired-end 150), with 1.2 million reads per library.

## Data Availability

Genomic data are available on NCBI Sequence Read Archive (SRA), GenBank, and GISAID (see accession numbers in Supplementary Table 2). All other data are included in the manuscript and the supplementary files.

## Supplementary data

**Supplementary Table 1:** Human monkeypox virus primer scheme (v1). Primer positions are determined based on alignment to the MT903345 reference genome.

**Supplementary Table 2:** Source data for samples included in this study. Listed are institute, specimen code, sample type, Ct value, sequencing platform, percent genome coverage at 10X, and accession numbers.

**Supplementary Figure 1:**
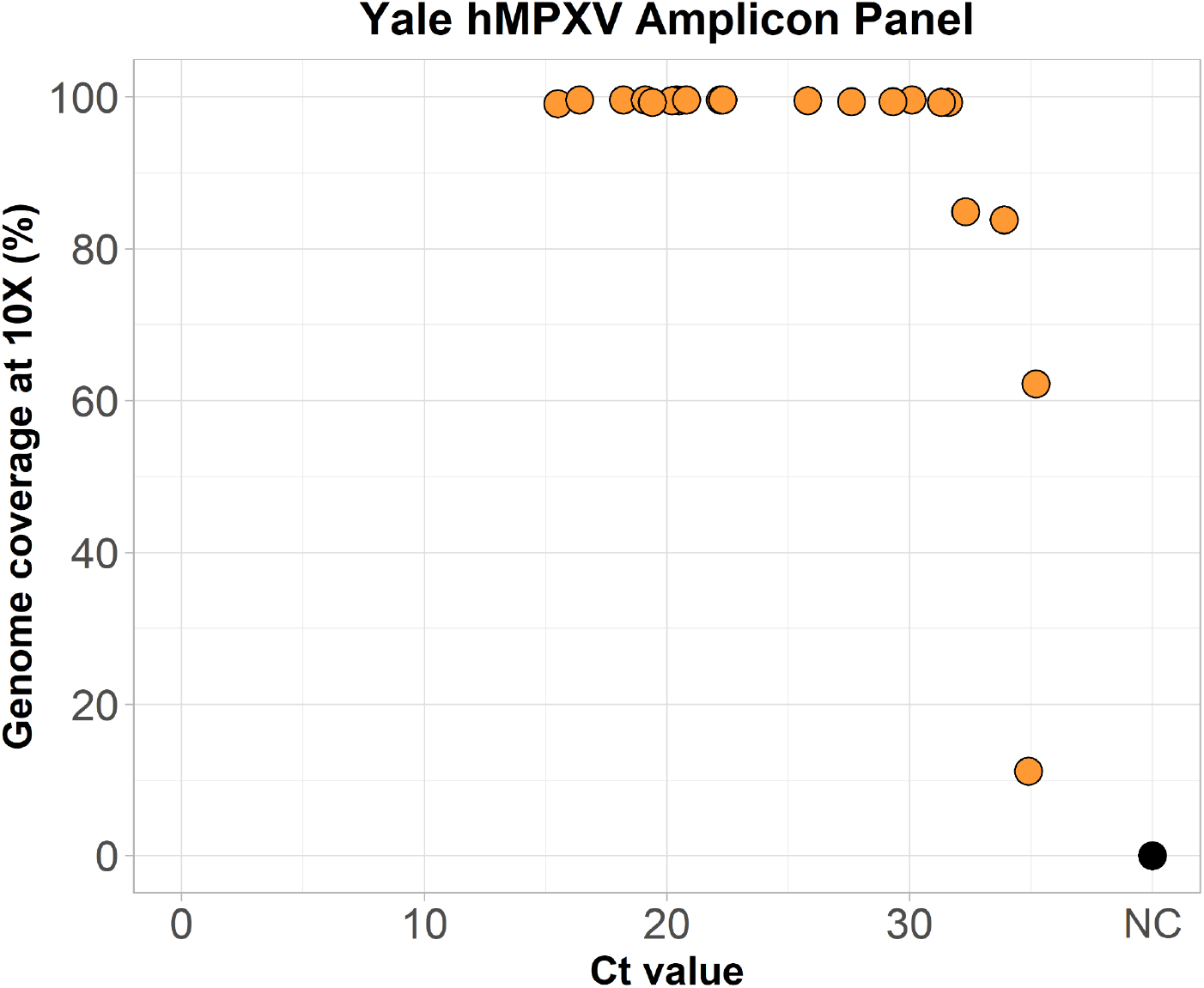
Validation of pre-pooled primers (Yale hMPXV amplicon panel) created by Integrated DNA Technologies. Lesion swabs (N=21) were re-sequenced using the Yale hMPXV amplicon panel instead of manually pooled primers. NC = negative control.

## Acknowledgements

We thank Cornelius Roemer for help with the logistic function analysis. This publication was made possible by CTSA Grant Number UL1 TR001863 from the National Center for Advancing Translational Science (NCATS), a component of the National Institutes of Health (NIH) awarded to CBFV. Its contents are solely the responsibility of the authors and do not necessarily represent the official views of NIH. INSA was partially funded by the HERA project (Grant/2021/PHF/23776) supported by the European Commission through the European Centre for Disease Control. The funders had no role in study design, data collection and analysis, decision to publish, or preparation of the manuscript.

## Data availability

Genomic data are available on NCBI Sequence Read Archive (SRA), GenBank, and GISAID (see accession numbers in **Supplementary Table 2**). All other data are included in the manuscript and the supplementary files.

## Author contributions

NFGC, CC, LG, GRG, and CBFV designed the study; all authors contributed to data/sample collection, sequencing, or bioinformatic analysis; NFGC, CC, LG, MD, DJP, NDG, GRG, CBFV analyzed the data; NFGC, CC, and CBFV drafted the manuscript; all authors reviewed and approved the manuscript.

