## Supplementary figures and images for "Development of an amplicon-based sequencing approach in response to the global emergence of human monkeypox virus"

### Supplementary Figure 1

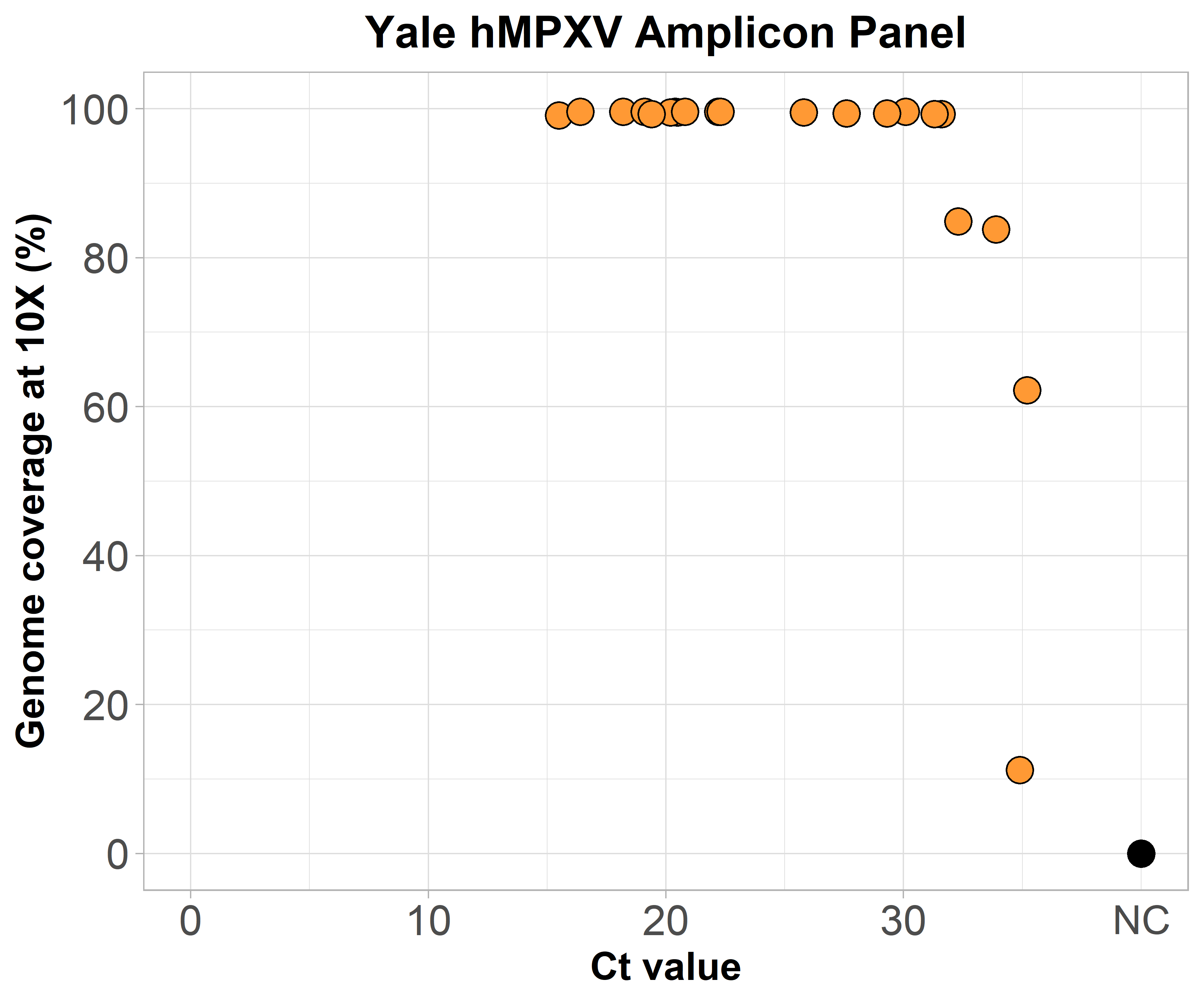
